# Brooding and neuroticism are strongly interrelated manifestations of the phenome of depression

**DOI:** 10.1101/2023.05.17.23290082

**Authors:** Asara Vasupanrajit, Ketsupar Jirakran, Chavit Tunvirachaisakul, Michael Maes

## Abstract

Neuroticism is a subclinical manifestation of the phenome of depression, comprising depressive and anxiety symptoms, and suicidal behaviors. Rumination is positively associated with depression and neuroticism and may mediate the effects of neuroticism on depression. This study aims to determine whether rumination or its components, including brooding or reflection, mediate the effects of neuroticism on depression, or alternatively, whether both neuroticism and rumination are manifestations of the phenome of depression. This study recruited 74 depressed subjects and 44 healthy controls. The depression group was split into groups with high versus low brooding scores. We used partial least squares (PLS) to examine mediation effects. We found that brooding and reflection scores are significantly higher in depressed patients than in controls. Patients with higher brooding scores have increased severity of depression, anxiety, insomnia, neuroticism, and current suicidal ideation as compared with patients with lower brooding scores and controls. There is a strong positive association between rumination, and neuroticism, depression, anxiety, and lifetime and current suicidal behaviors. PLS analysis shows that brooding does not mediate the effects of neuroticism on the depression phenome, because no discriminant validity could be established between neuroticism and brooding, or between neuroticism and brooding and the depression phenome. We were able to extract one validated latent vector from brooding and neuroticism, insomnia, depression, anxiety, and current suicidal behaviors. Overall, this study supports the theory that rumination and neuroticism are manifestations of the phenome of depression, just like affective symptoms, suicidal behaviors, and insomnia.

## Introduction

Depression is one of the most common mental disorders and a significant global burden (Liu et al., 2020; Rehm and Shield, 2019; Vos et al., 2020). With an average onset age of 24 years, the 12-month and lifetime prevalence of depression are approximately 5.9% and 11.1%, respectively (Bromet et al., 2011). Individuals with depression exhibit a variety of symptoms, including loss of interest, sadness, anxious mood, inability to concentrate, sleep problems, low self-esteem, and suicidal ideation or behaviors (American Psychiatric Association, 2013; Malhi and Mann, 2018; World Health Organization, 2004). Recent evidence suggests that neuroticism, a personality trait, increases the risk of depression (Jirakran et al., 2023; Vittengl, 2017). A common psychological explanation is that the chronic negative emotional trait associated with neuroticism may result in an aberrant response to environmental stressors, thereby increasing the likelihood of hopelessness and, consequently, depression (Widiger and Oltmanns, 2017). Moreover, a number of studies demonstrate that depressive patients have cognitive impairments in attention and working memory that result in a selective focus on negative stimuli (De Raedt and Koster, 2010; Koster et al., 2011; Semkovska et al., 2019; Whitmer and Banich, 2007) and exposure to unavoidable stressors that activate depressogenic thoughts (Andrews and Thomson, 2009; De Raedt and Koster, 2010; Koster et al., 2011).

Rumination, a cognitive process characterized by a pattern of activities and/or thoughts that inhibit mood and impede problem-solving, instrumental activity, and social support, frequently accompanies depression (Nolen-Hoeksema, 1991, 2000; Nolen-Hoeksema et al., 2008). Rumination consists of two factors: brooding and reflection (Treynor et al., 2003). In contrast to brooding, which consists of dwelling on unpleasant feelings over and over again, reflective thinking comprises active cognitive problem solving or adaptive thinking, with the goal of alleviating such feelings (Treynor et al., 2003).

Earlier research showed a substantial link between ruminating and neuroticism (Riihimäki et al., 2016). Moreover, ruminating, especially brooding, may contribute to the onset of depression and predict suicidal behaviors (Chiang et al., 2022; Hosseinichimeh et al., 2018; Riihimäki et al., 2016; Tang et al., 2021), and may mediate the effects of neuroticism on depression (Lyon et al., 2021; Roelofs et al., 2008; Vidal-Arenas et al., 2022). Andrews and Thomson (2009) have proposed that depressed patients show the tendency to repeat their thoughts and analyze their problems, thereby disrupting concentration processes and inducing anhedonia and depression. Rumination may impede working memory processes by retaining outdated knowledge and focusing on unpleasant information, resulting in memory errors that may function as long-term stressors (Bruning et al., 2023). In addition, rumination is reported to mediate the effects of depressive mood on sleep quality, suggesting that a psychological mechanism mediates the effects on insomnia. Pre-sleep rumination also predicts poor sleep quality, including delayed sleep onset in individuals with depressive symptoms (Pillai et al., 2014).

Recent research has shown, however, that a latent vector may be extracted from neuroticism (a trait) and the acute phenome (symptomatology of severe depression), which consists of the severity of depression and anxiety, as well as suicidal thoughts (SI) and attempts (SA) (Jirakran et al., 2023; Maes et al., 2023b). This suggests that neuroticism is a subclinical expression of the phenome of major depressive disorder (Jirakran et al., 2023; Maes et al., 2023b). Nevertheless, no studies have examined whether ruminating may mediate the effects of neuroticism on the phenome of depression, including suicidal behaviors and sleep disorders, or alternatively, that rumination is a feature of the phenome of depression.

Hence, the aims of the present study are to determine whether a) rumination mediates the effects of neuroticism on the phenome of depression, including suicidal behaviors and sleep disorders; or b) rumination and neuroticism are, as depression, anxiety, insomnia, and suicidal ideation, manifestations of the depression phenome. Toward this end, we use partial least squares (PLS) analysis, which combines regression, mediation, and factor analysis.

## Material and methods

### Participants

Participants were recruited from the outpatients Department of Psychiatry, King Chulalongkorn Memorial Hospital, Bangkok, Thailand between November 2021 to February 2023. This research recruited Thai-speaking undergraduate or graduate students aged between 18 and 35, of both sexes, and studying at any university faculty, Bangkok. The students were diagnosed by a senior psychiatrist as suffering from depressive disorders using DSM-5 (American Psychiatric Association, 2013) and ICD-10 (World Health Organization, 2004) criteria; codes F32 (depressive episode), F33 (major depressive disorder, recurrent), and F34.1 (dysthymic disorder). Furthermore, they had a Hamilton Depression Rating Scale (HAM-D) (Hamilton, 1960) score ≥7. The patient group was divided into high and low brooding groups using the median-split method. Exclusion criteria were: other axis 1 psychiatric disorders, including schizophrenia, alcohol or substance use disorders, psycho-organic disorders, anxiety disorders, autism, bipolar disorder, schizoaffective disorder; individuals with high risk to committing suicide; current medical illness including immune and neuro-inflammatory disorders, such as diabetes type 1, chronic kidney disease, endocrine or autoimmune disorders, psoriasis, lupus erythematosus, Parkinson’s disease, Alzheimer’s disease, multiple sclerosis, and stroke. Pregnant and lactating female students were excluded to participate. Healthy controls were included if their lifetime history of psychiatric disorders and suicidality was negative and the HAM-D (Hamilton, 1960) score < 7. They were recruited through word of mouth and online advertisements and were matched to the patients for age, sex, and number of education years. A total of 118 students were included in this study. This is a case-control study reviewed and approved by the Institutional Review Board (IRB) of the Faculty of Medicine, Chulalongkorn University, Bangkok, Thailand (IRB No.351/63). All participants gave written informed consent prior to the study.

### Measures

We used a semi-structured interview to obtain socio-demographic data comprising age, sex, relationship status, year of education, current smoking, lifetime history of COVID-19, family history of mental health, including major depressive disorder, bipolar disorder, anxiety, psychotic and suicidal histories. The Ruminative Response Scale (RRS) (Nolen-Hoeksema and Morrow, 1991) has been used to assess rumination. The RRS comprises 22 items rated on a 4-points scale ranging from almost never (1) to almost always (4). This study used the Thai version (Thanoi et al., 2011) which shows good internal consistency (α = 0.90) and content validity (CVI= 0.95). The International Personality Item Pool-NEO (IPIP-NEO) (Socha et al., 2010) was developed based on the Big Five personality traits, and comprises 50 phrases describing an individual’s behavior, which were selected as proxies for the broad domain scores of NEO-PI-R (Costa and McCrae, 1992). This study used the Thai version translated by Yomaboot and Cooper (2016). The Thai version was examined using exploratory and confirmatory factor analysis, which revealed an acceptable fit and a five-factor model consisting of eight items for neuroticism (N), three items for extraversion (E), five items for agreeableness (A), six items for openness (O), and eight items for consciousness (C). Internal consistency for this 30-item version was acceptable to good (Cronbach’s alpha for N = 0.83, E = 0.76, A = 0.37, O = 0.67, C = 0.73).

This study used the HAM-D (Hamilton, 1960) and the Beck Depression Inventory-II (BDI-II) (Beck et al., 1996) to evaluate the severity of depression. We used the HAM-D Thai version, translated by Lotrakul et al. (1996). The BDI-II (Beck et al., 1996) is a self-rating questionnaire comprising 21 items rated on a 4-points scale ranging from not present (0) to severe (3). The Thai version of the BDI-II was translated by Mungpanich (2008) and has excellent validity and reliability. The State-Trait Anxiety Inventory (STAI) (Spielberger et al., 1983) is a self-rating questionnaire to assess state anxiety and comprises 20 items ranging from 1 (not at all) to 4 (mostly). The total score ranges from 20 to 80 which higher scores indicating greater anxiety. The Thai version was translated by Iamsupasit and Phumivuthisarn (2005) and has good internal consistency with a Cronbach’s alpha coefficient ranging from 0.86 to 0.92.The Columbia–Suicide Severity Rating Scale (C-SSRS) (Posner et al., 2011) is a semi-structured interview that was used to evaluate lifetime (until one moth prior to inclusion of the participants) and current (within 1 month) suicidal behaviors (SB). The C-SSRS is divided into four constructs including severity of suicidal ideation, intensity of suicidal ideation, suicidal behaviors (self-harm and attempts), and the lethality subscale. The Thai version was provided by The Columbia Lighthouse Project (2016).

The Insomnia Severity Index (ISI) (Morin, 1993; Morin et al., 2011) is a self-rating questionnaire to assess severity of sleep problems. The ISI comprises 7 Likert-scale items ranging from 0 (no problem) to 4 (very severe problems). The total score of 0-7 is interpreted as the absence of insomnia; 8-14 as sub-threshold insomnia; 15-21 as moderate insomnia; and 22-28 as severe insomnia. The ISI Thai version is provided by the Mapi Research Trust (Mapi Language Services, 2015). The body mass index (BMI), BMI was computed as weight (kg) divided by height (m) squared.

### Statistical analyses

Using Pearson’s product-moment correlation coefficients, the analysis investigated the correlations between continuous variables. Analysis of variance (ANOVA) was utilized to investigate the associations between diagnostic groups and clinical data. A contingency table analysis (χ2-test) was used to evaluate statistical associations between categorical variables. Principal component analysis (PCA) was utilized to reduce the number of items into a single PC score, which could then be utilized in other statistical analyses. Using the Kaiser-Meyer-Olkin (KMO) test for sample adequacy, which is considered adequate when > 0.6, and the Bartlett’s sphericity test, factorability was evaluated. The first PC is admitted only if the variance explained (VE) is greater than 50 percent and all loadings on the first PC are greater than 0.7. Using SmartPLS path analysis (Ringle et al., 2015), we evaluated the causal relationships between neuroticism, rumination (brooding and reflection), and phenome of depression. The latter was conceptualized as a latent vector extracted from the severity of depression (HAMD, BDI-II), anxiety (STAI), insomnia (ISI score), and current SB. Neuroticism was conceptualized as a latent vector extracted from 6 IPIP-NEO items, brooding as a latent vector extracted from 14 RRS, and reflection as a latent vector extracted from 3 RRS items. Other indicators were entered as singular indicator (age and sex). Complete PLS analysis was performed when the outer and inner models satisfied predefined quality criteria, namely: a) all loadings on the latent vectors are greater than 0.7 at p < 0.001; b) the model fit SRMR is less than 0.08; c) all latent vectors have adequate composite reliability (> 0.7), Cronbach’s alpha (> 0.7), and rho A (> 0.8) scores, with an average variance extracted (AVE) greater than 0.50; d) the models are not incorrectly specified as reflective models, as confirmed by Confirmatory Tetrad Analysis. Discriminant validity of the constructs was checked using the Heterotrait-Monotrait ratio (HTMT) with a cut-off value of 0.85. Using 5,000 bootstrap samples, a complete PLS analysis was conducted to calculate specific indirect, total indirect, and total direct path coefficients (with exact p-values).

## Results

### Results of PC analysis

We were not able to extract one validated PC from all RRS items. However, we were able to extract one validated PC from items 1, 2, 3, 4, 5, 6, 9, 10, 13, 14, 15, 16, 17, 18, 19, and 22, as shown in **Table 1**. Since, all items on this validated PC score highly on brooding items, it was labeled as “PC_brooding”. **Electronic Supplementary File (ESF), Table 1** lists all RRS items. We were also able to extract one PC from items 7, 11, 20, and 21, as shown in Table 1. Since these 4 items score highly on self-reflection, it was labeled as “PC_reflection”. Using PC_brooding and PC_reflection, we constructed a composite, which is a weighted composite of rumination.

**Table 1.** Results of principal component (PC) analysis and construction of brooding and reflection principal components (PCs) based on the items of the Ruminative Response Scale (RRS) rating scale.

| **PC_brooding** |  | **PC-reflection** |  |
| --- | --- | --- | --- |
| **Variables** | **Loading** | **Variables** | **Loading** |
| RRS1 | 0.791 | RRS7 | 0.851 |
| RRS2 | 0.754 | RRS11 | 0.788 |
| RRS3 | 0.842 | RRS20 | 0.856 |
| RRS4 | 0.793 | RRS21 | 0.701 |
| RRS5 | 0.722 |  |  |
| RRS6 | 0.771 |  |  |
| RRS9 | 0.762 |  |  |
| RRS10 | 0.567 |  |  |
| RRS13 | 0.611 |  |  |
| RRS14 | 0.720 |  |  |
| RRS15 | 0.731 |  |  |
| RRS16 | 0.811 |  |  |
| RRS17 | 0.854 |  |  |
| RRS18 | 0.846 |  |  |
| RRS19 | 0.815 |  |  |
| RRS22 | 0.733 |  |  |
| KMO = 0.933 |  | KMO = 0.720 |  |
| $X^2 = 1357.987$ (df=120), $p < 0.001^{***}$ | | $X^2 = 176.048$ (df=6), $p < 0.001^{***}$ | |
| VE = 58.01% |  | VE = 64.27% |  |

KMO: The Kaiser-Meyer-Olkin Test; VE: Variance explained; $X^2$ : Bartlett's test of sphericity. ESF, table 1 shows the explanation of the RRS items

In accordance with previous research (Jirakran et al., 2023; Maes et al., 2021), we made new constructs reflecting lifetime (LT) and current SB, including SI and SA. **ESF, Table 2** and **ESF, Table 3** show how we constructed LT_SI, LT_SA, LT_SB, current_SI, current_SA, and current_SB. To make a new comprehensive phenome index, we extracted the first PC from the total HAM-D (loading=0.918), BDI-II (loading=0.922), STAI (loading=0.745), ISI (loading=0.767), and current_SB (loading=0.758) scores (KMO=0.830, Bartlett’s χ2=346.103, df=10, p<0.001, explained variance=68.229%). This validated PC was labeled “current_phenome”.

### Socio-demographic and clinical data

To display differences in depression patients with and without high brooding scores, we have binned the patient group into two subgroups based on the median values of PC_brooding. **Table 2** shows the socio-demographic and basic clinical features of the participants. There were no significant differences in age, sex, BMI, number of education years, current smoking frequency between depression with high and low PC_brooding scores and controls. There are no significant differences in depression ICD10 subtypes, duration of depression, medication status, lifetime of COVID-19 infection, and psychiatric and suicide history among family members between both patient groups.

**Table 2.** Demographic and clinical data of depressive patients with high versus low brooding scores, and healthy controls (HC)

| Variables | HC<br>(n=44) | Depression-low<br>brooding<br>(n=37) | Depression-high brooding<br>(n=37) | F/$\chi^2$<br>/FET | df | p |
| --- | --- | --- | --- | --- | --- | --- |
| Age (years) | 23.5 (3.2) | 22.5 (3.5) | 22.2 (2.4) | 1.99 | 2/115 | 0.141 |
| Sex (Male/Female) | 7/37 | 6/31 | 6/31 | 0.00 | 2 | 0.999 |
| Education (years) | 16.6 (1.5) | 16.1 (3.0) | 15.5 (2.2) | 2.57 | 2/115 | 0.081 |
| BMI (kg/m<sup>2</sup>) | 22.3 (3.6) | 22.4 (4.8) | 22.1 (5.4) | 0.03 | 2/115 | 0.966 |
| Relationship status (single/boy or girlfriends) | 27/17 | 16/21 | 14/23 | 7.27 | 2 | 0.122 |
| Current smoking (yes/no) | 1/43 | 3/34 | 3/34 | 1.84 | 2 | 0.432 |
| Lifetime Covid-19 history (yes/no) | 14/30 <sup>c</sup> | 12/25 | 4/33 <sup>a</sup> | 6.07 | 2 | 0.048 |
| ICD-10 Diagnosis (F32/F33/F34) | - | 34/0/3 | 33/3/1 | 4.02 | 2 | 0.134 |
| Duration of depression (months) | - | 21.0 (16.6) | 27.6 (26.7) | 1.63 | 1/72 | 0.206 |
| Psychiatric medication |  |  |  |  |  |  |
| Antidepressant (yes/no) | - | 34/3 | 36/1 | FET | - | 0.615 |
| Benzodiazepine (yes/no) | - | 15/22 | 22/15 | 2.65 | - | 0.104 |
| Antipsychotic (yes/no) | - | 7/30 | 7/30 | 0.00 | 1 | 1.000 |
| Mood stabilizer (yes/no) | - | 3/34 | 2/35 | FET | - | 1.000 |
| Psychiatric history in family |  |  |  |  |  |  |
| Depression (yes/no) | - | 7/30 | 5/32 | 0.40 | - | 0.754 |
| Bipolar (yes/no) | - | 1/36 | 0/37 | FET | - | 1.000 |
| Anxiety (yes/no) | - | 2/35 | 1/36 | FET | - | 1.000 |
| Psychotic (yes/no) | - | 1/36 | 0/37 | FET | - | 1.000 |
| Suicide history in family (yes/no) | - | 4/33 | 3/34 | FET | - | 1.000 |
| Total RRS score | 44.2 (12.4) <sup>b,c</sup> | 59.6 (7.6) <sup>a,c</sup> | 75.6 (6.7) <sup>a,b</sup> | 110.81 | 2/115 | <0.001 |
| PC_brooding | -0.888 (0.796) <sup>b,c</sup> | -0.024 (0.502) <sup>a,c</sup> | 1.080 (0.278) <sup>a,b</sup> | 114.47 | 2/115 | <0.001 |
| PC_reflection | -0.817 (0.838) <sup>b,c</sup> | 0.270 (0.675) <sup>a,c</sup> | 0.701 (0.747) <sup>a,b</sup> | 43.33 | 2/115 | <0.001 |

Results are shown as mean (S.D.); F: results of analysis of variance; $\chi^2$ : analysis of contingency tables; FET: Fisher-exact probability test; RRS: The Ruminative Response Scale; PC: Principal component; <sup>a, b, c</sup> pairwise comparisons using LSD (p<0.05), i.e. <sup>a</sup> means significant different from HC, <sup>b</sup> means significant different from Depression-low brooding, <sup>c</sup> means significant different from Depression-high brooding.

### Clinical features of patients with current suicidal ideation

**Table 3** shows the clinical features of healthy controls and both patient groups. The HAM-D, BDI-II, STAI, ISI and current phenome scores were significantly different between the three study groups and increased from controls → low brooding → high brooding groups. LT and current SI, SA, and SB were significantly higher in both patient groups than controls. Nevertheless, current_SI was significantly different between the three study groups with the highest values in those with high brooding. Furthermore, there was a significant difference in three dimensions of personality, namely higher neuroticism, lower agreeableness, and lower conscientiousness, in both depressive groups as compared with controls. The most significant personality trait associated with depression was neuroticism. In analogy with Jirakran et al. (2023), this study will, therefore, focus on neuroticism.

**Table 3.**
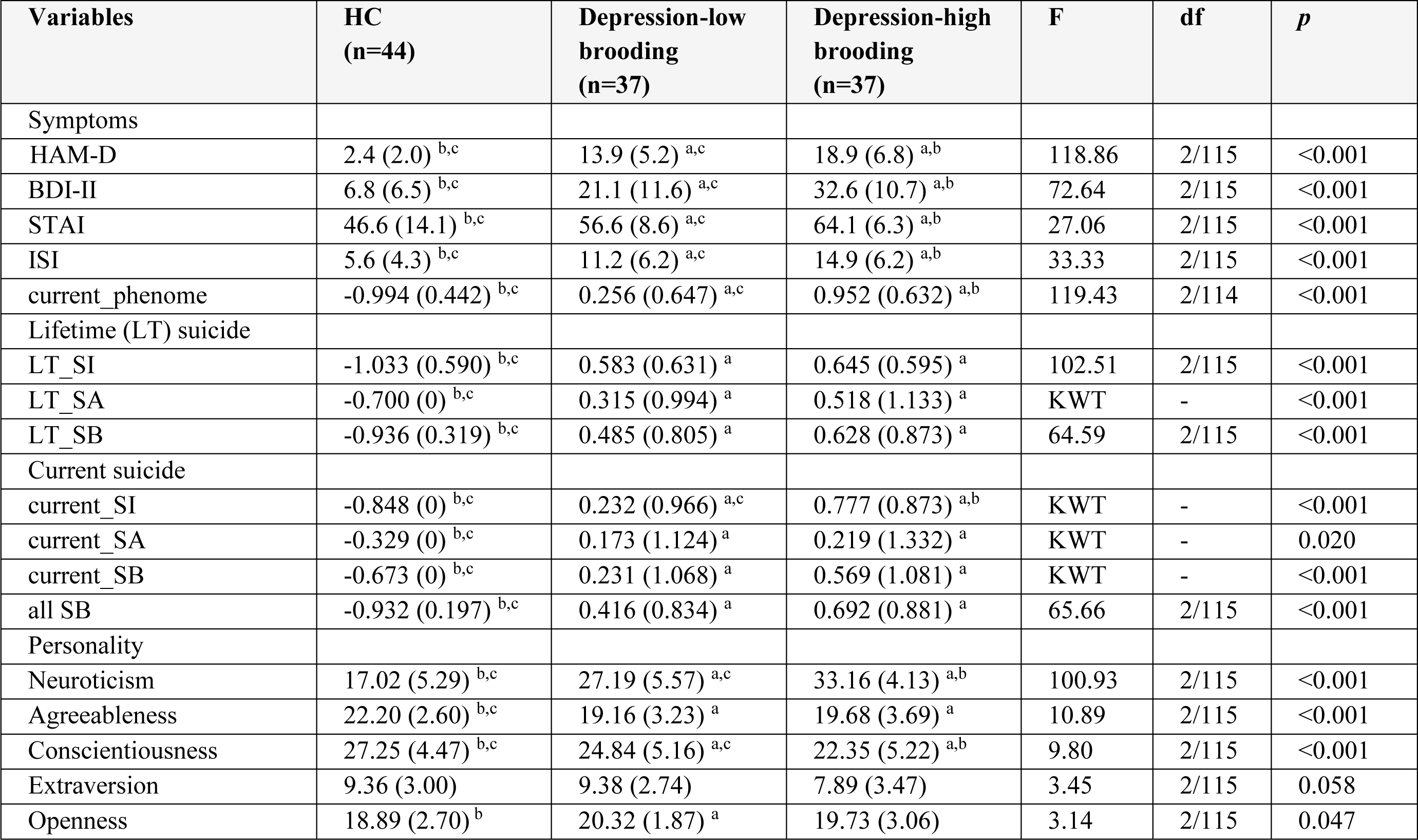
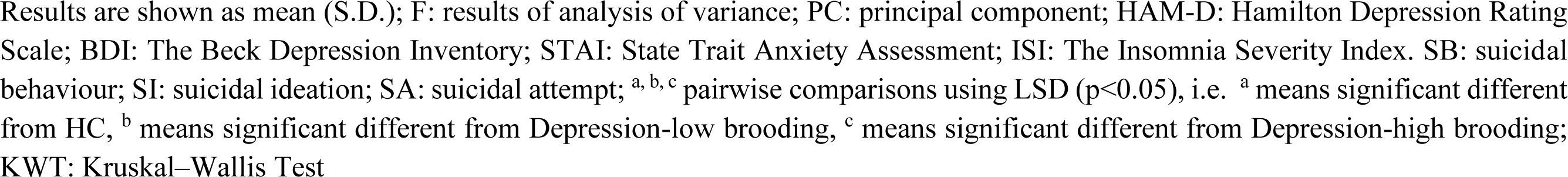
Features of depressive patients with high versus low brooding scores and healthy controls (HC)

### Intercorrelation matrix

The correlations between rumination and clinical data, including current phenome, HAM-D, BDI-II, STAI, ISI, LT and current SI, SA, SB, and PC_neuroticism are shown in **Table 4**. There were positive relationships between rumination, as well as PC_brooding, and all clinical variables (*r*=0.4-0.8, p≤0.001). Rumination, both brooding and reflection, showed strong positive associations with the current phenome and neuroticism. **Figure 1** shows the partial regression of current_phenome on PC_brooding (after controlling for the effects of age, sex, and education). **Figure 2** shows the partial regression of neuroticism on PC_brooding and **Figure 3** shows the partial regression of current_phenome on neuroticism. There was a significant correlation between rumination and PC_brooding, and LT and current SA. PC_reflection showed positive relationships with all clinical variables, except current-SA.

**Figure 1.**
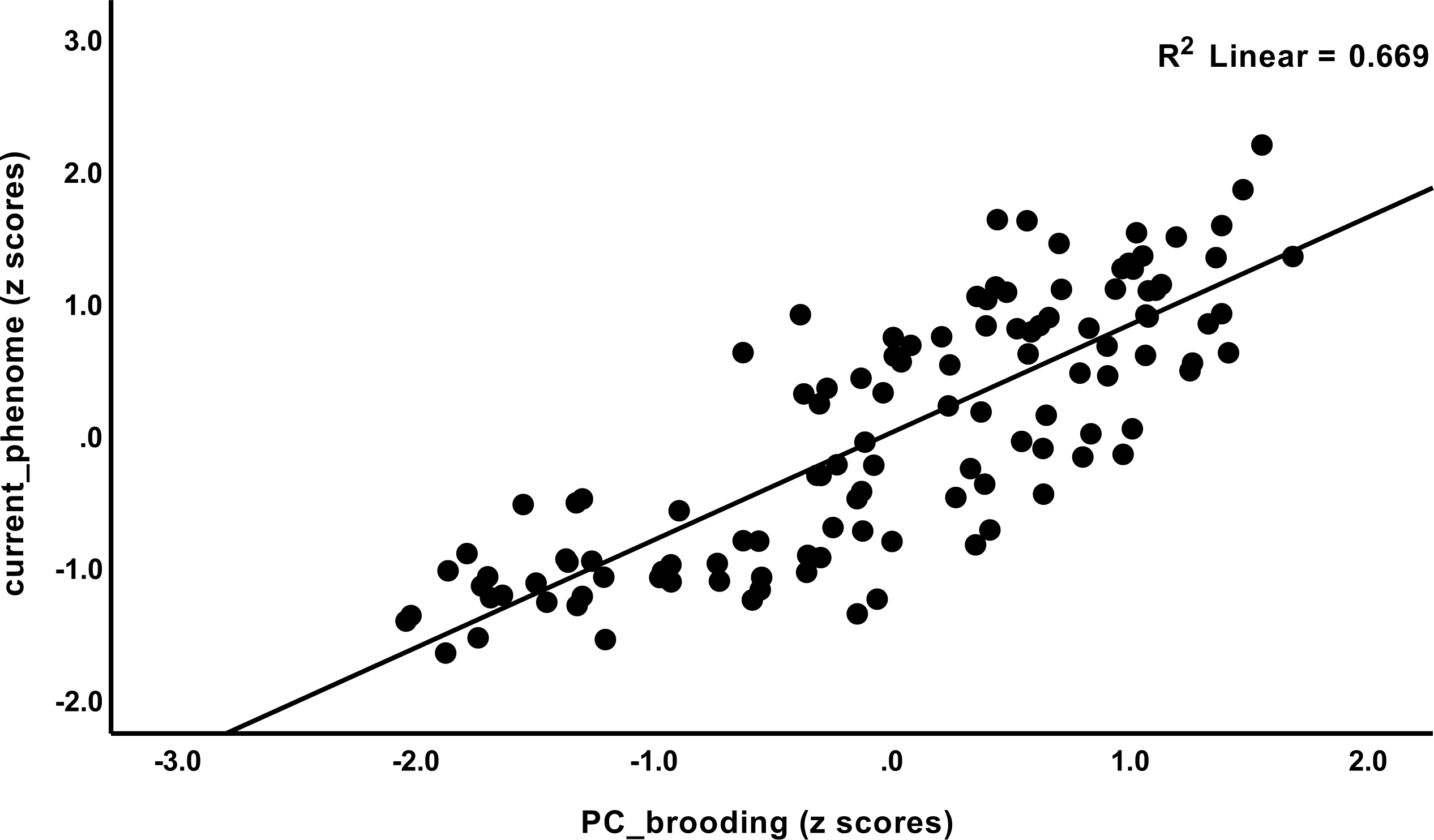
Partial regression of current_phenome on PC_brooding (after allowing for the effects of age, sex, and education).

**Figure 2.**
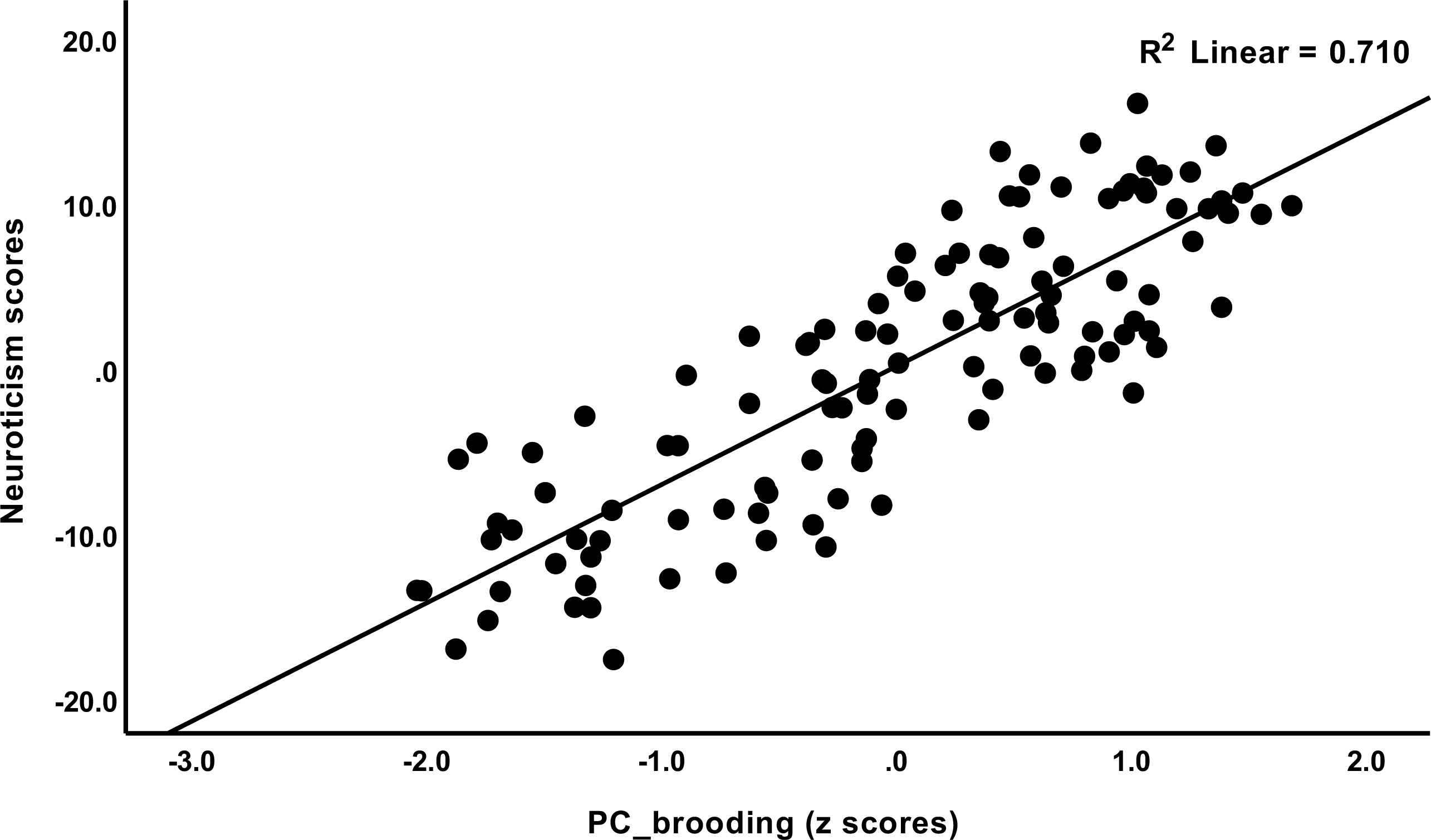
Partial regression of neuroticism on PC_brooding.

**Figure 3.**
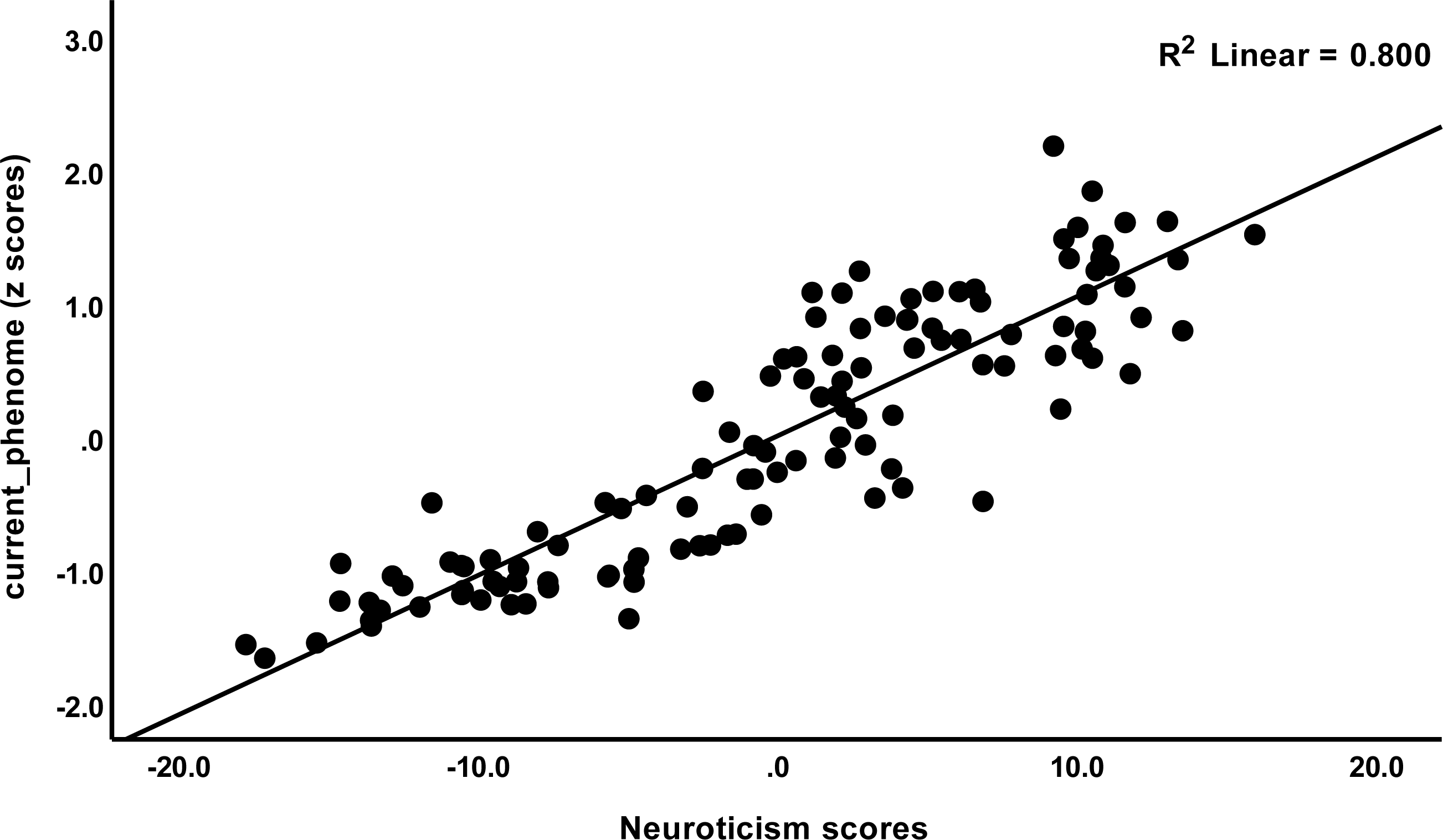
Partial regression of current_phenome on neuroticism.

**Table 4.** Correlation matrix between rumination, brooding and reflection and other clinical features of depression.

| **Variables** | **Rumination composite** | **PC_brooding** | **PC_reflection** |
| --- | --- | --- | --- |
| Rumination | 1 | - | - |
| PC_brooding | 0.921 (<0.001) | 1 | - |
| PC_reflection | 0.921 (<0.001) | 0.696 (<0.001) | 1 |
| current_phenome | 0.790 (<0.001) | 0.826 (<0.001) | 0.629 (<0.001) |
| HAM-D | 0.705 (<0.001) | 0.706 (<0.001) | 0.593 (<0.001) |
| BDI-II | 0.733 (<0.001) | 0.771 (<0.001) | 0.578 (<0.001) |
| STAI | 0.656 (<0.001) | 0.761 (<0.001) | 0.447 (<0.001) |
| ISI | 0.632 (<0.001) | 0.605 (<0.001) | 0.559 (<0.001) |
| Lifetime (LT) suicide |  |  |  |
| LT_SI | 0.666 (<0.001) | 0.647 (<0.001) | 0.579 (<0.001) |
| LT_SA | 0.392 (<0.001) | 0.399 (<0.001) | 0.324 (<0.001) |
| LT_SB | 0.572 (<0.001) | 0.565 (<0.001) | 0.488 (<0.001) |
| Current suicide |  |  |  |
| current_SI | 0.572 (0.001) | 0.594 (<0.001) | 0.460 (<0.001) |
| current_SA | 0.199 (0.030) | 0.242 (0.008) | 0.125 (0.179) |
| current_SB | 0.790 (<0.001) | 0.478 (<0.001) | 0.334 (<0.001) |
| all SB | 0.587 (<0.001) | 0.601 (<0.001) | 0.480 (<0.001) |
| Neuroticism | 0.870 (<0.001) | 0.837 (<0.001) | 0.517 (<0.001) |

Results of Spearman's rank-order correlation calculations; PC: principal components; HAM-D: Hamilton Depression Rating Scale; BDI: The Beck Depression Inventory; STAI: State Trait Anxiety Assessment; ISI: The Insomnia Severity Index; SB: suicidal behaviour; SI: suicidal ideation; SA: suicidal attempt; RRS: The Ruminative Response Scale.

### Results from PLS analysis

Based on the theories described in the Introduction, we constructed a first PLS model as shown in **Figure 4**. The depression phenome was conceptualized as a latent vector extracted from HAM-D, BDI-II, STAI, ISI, and current-SB scores. Neuroticism was considered to be a latent vector extracted from six IPIP-NEO neuroticism subdomain items. We also constructed brooding and reflection latent vectors and entered the items shown in Table 2 as indicators. The neuroticism, brooding and reflection latent vectors predicted the phenome, and brooding could mediate the effects of neuroticism on the phenome. According to psychological theories that distress, and brooding may lead to reflection, PC_brooding was related with PC_reflection. Age was employed as a single explanatory variable that could predict all latent vectors. The model in Figure 4 shows only the significant paths. The model fit quality was adequate with SRMR = 0.066. Neuroticism, brooding, reflection, and the phenome revealed adequate construct validity and convergence with a) an average variance extracted (AVEs) of 0.769, 0.617, 0.730, and 0.680, respectively; b) Cronbach’s alpha of 0.940, 0.952, 0.817, and 0.880, respectively; and c) composite reliability of 0.952, 0.957, 0.890, and 0.913, respectively. All loadings of the outer model were greater than 0.7. We found that 79.3% of the phenome variance was explained by neuroticism and brooding. There were specific indirect effects of neuroticism on the phenome (t=5.26, p<0.001) and on reflection (t=11.63, p<0.001), which were mediated by brooding. Age showed a specific indirect effect on the phenome that was mediated by neuroticism (t=2.03, p=0.043). There were significant total effects of neuroticism (t=37.19, p<0.001) and brooding (t=5.25, p<0.001) on the phenome, whereas age showed an inverse total effect (t=-2.13, p=0.033).

**Figure 4.**
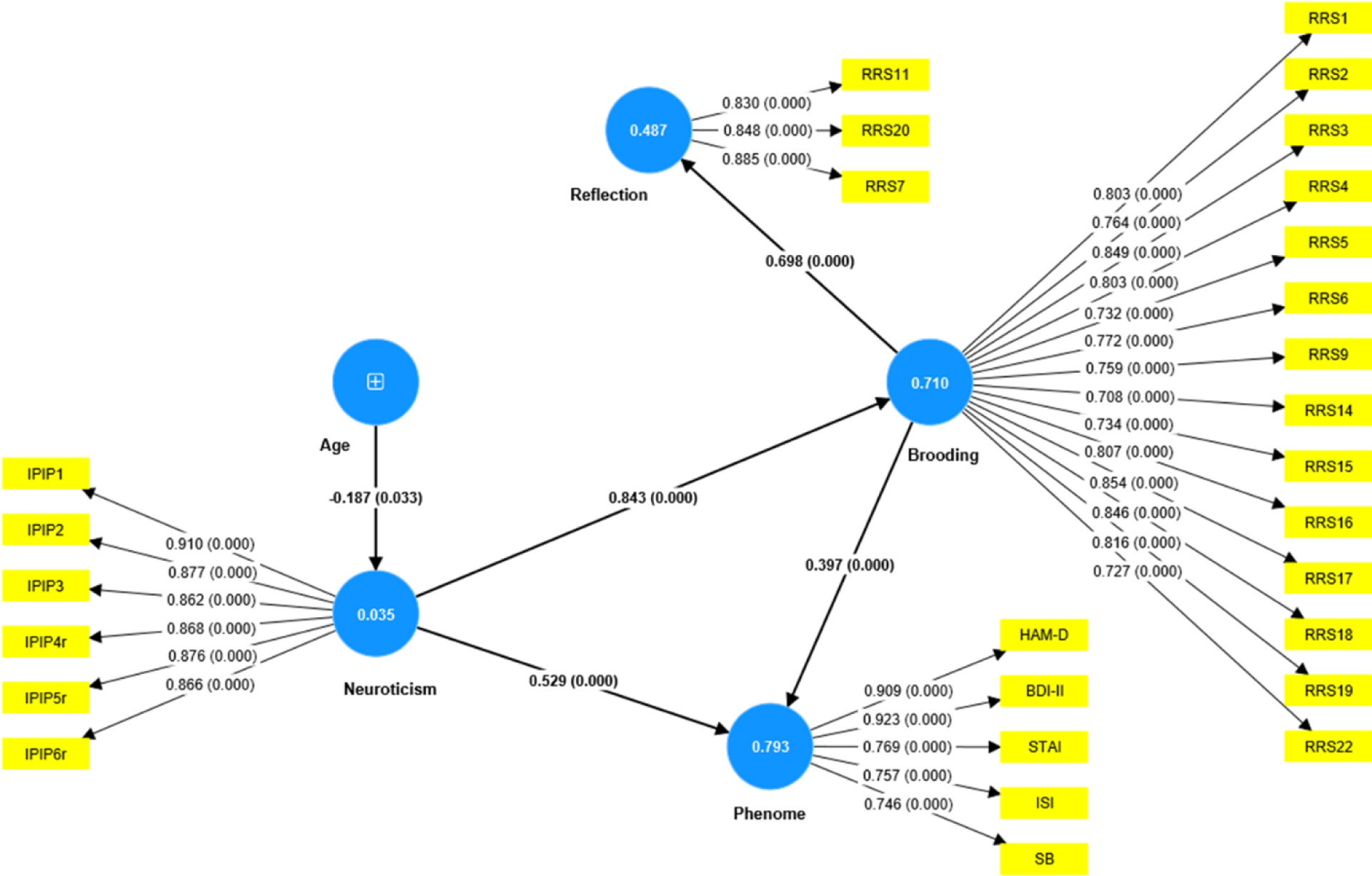
Results of PLS analysis. Shown are the significant paths, including the path coefficients (with exact p value) of the inner model, and loadings (with p values) of the outer model. Figures in blue circles indicate explained variance. Age was entered as a single indicator (denoted as +); neuroticism, phenome, brooding, and reflection were entered as latent vectors extracted from their manifestations. IPIP: The International Personality Item Pool-NEO (IPIP-NEO), HAM-D: Hamilton Depression Rating Scale; BDI-II: The Beck Depression Inventory; STAI: State Trait Anxiety Assessment; ISI: The Insomnia Severity Index; SB: suicidal behaviours; RRS: The Ruminative Response Scale.

Nevertheless, this first model is not adequate because no significant discriminatory validity could be established between the phenome and either neuroticism (HTMT=0.942) and brooding (HTMT=0.915), and between neuroticism and brooding (HTMT=0.884). Interestingly, discriminant validity could be established between reflection and brooding (HTMT=0.776), reflection and neuroticism (HTMT=0.669), and reflection and the phenome (HTMT=0.747). Therefore, we have tried to combine both neuroticism and brooding into the same phenome latent vector (see **Figure 5**.). Age was introduced to the model as an explanatory variable of the phenome. This model fit was adequate with SRMR = 0.051. The overall phenome latent vector showed adequate validity with an AVE of 0.672, Cronbach’s alpha of 0.919, and composite reliability of 0.971. All loadings were greater than 0.7. PC_reflection showed a loading of only 0.641 and thus could not be included.

**Figure 5.**
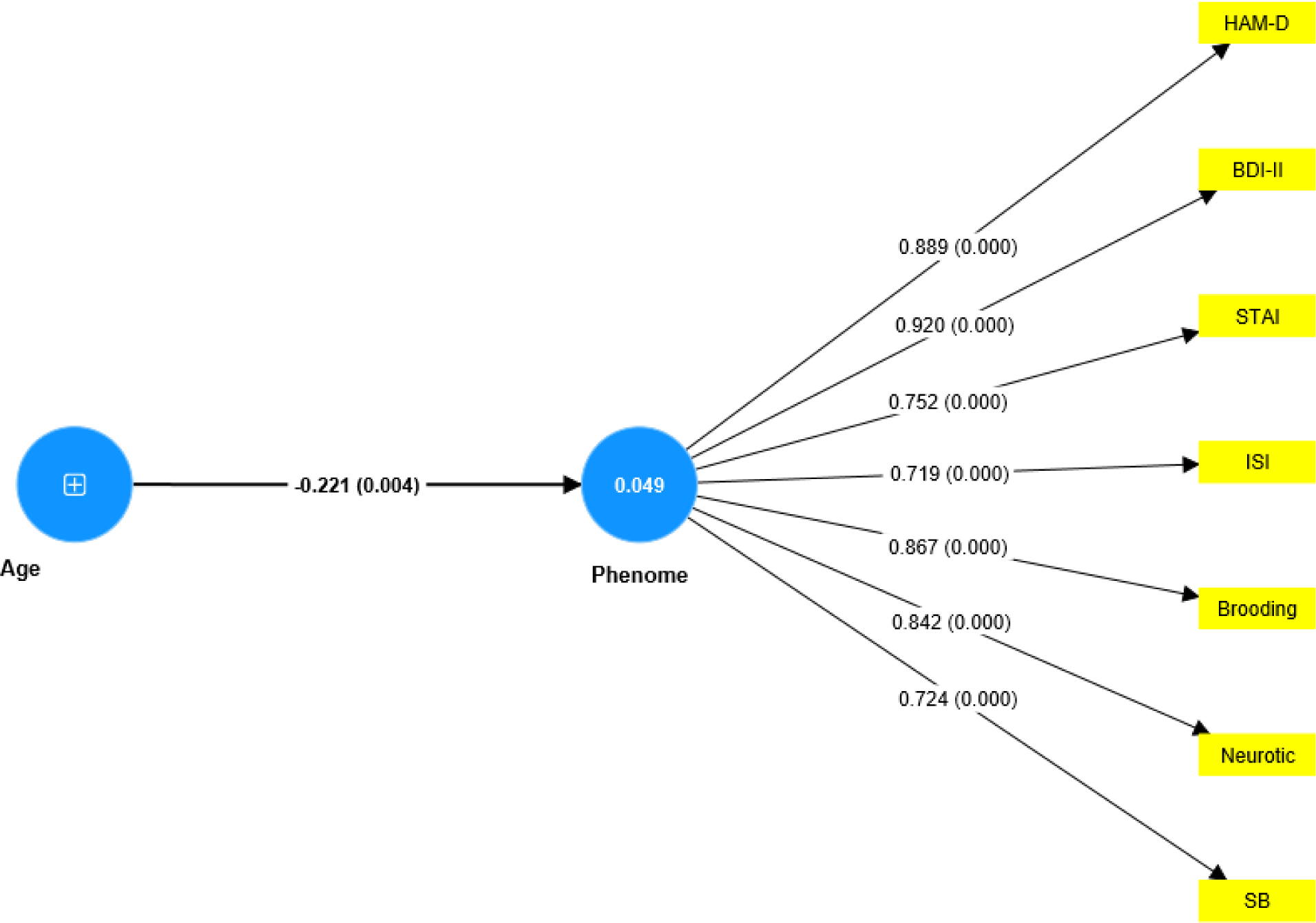
Results of PLS analysis. Shown are the significant paths, including the path coefficients (with exact p value) of the inner model, and loadings (with p values) of the outer model. Figures in blue circles indicate the explained variance. Age was entered as a single indicator (denoted as +); the Phenome was entered as a latent vector extracted from HAM-D: Hamilton Depression Rating Scale; BDI-II: The Beck Depression Inventory; STAI: State Trait Anxiety Assessment; ISI: The Insomnia Severity Index; SB: suicidal behaviour

## Discussion

### Increased brooding and reflection in depression

The first main finding of this study is that rumination scores, including both brooding and reflection, were significantly higher in depressed patients than in healthy controls, with brooding having a stronger association than reflection. According to Treynor et al. (2003), brooding is a maladaptive fixation on distress symptoms, whereas reflection is an adaptive way to engage in cognitive problem-solving. Previous research has shown that brooding is a significant predictor of depression over time, whereas there is less evidence that reflection has the same effect (Armey et al., 2009; Burwell and Shirk, 2007; Young et al., 2014). Previous research has shown that rumination and introspection are associated with the development of depressive symptoms, and that rumination may predict the onset of depression (Nolen-Hoeksema, 2000; Owens and Gibb, 2017; Young et al., 2014). Other studies demonstrated a correlation between rumination and the severity (Riihimäki et al., 2016), duration (Riihimäki et al., 2016), and recovery (Kuehner and Weber, 1999) of depression. According to Koster et al. (2011), rumination indicates a lack of cognitive control to divert attention away from negative self-references. Owens and Gibb (2017) found that ruminating was associated with enhanced selective attention to depression-related information, resulting in an increased risk of depression, onset, and relapse. It is possible that ruminations interfere with reward systems, causing depressed individuals to repeatedly focus on negative outcomes, such as negative self-evaluations and judgments, resulting in a diminished ability to generate and experience positive affects (Koster et al., 2011; Rutherford et al., 2023).

### Brooding as a symptom of depression

The second major finding of our study is that no discriminatory validity was established between rumination and the phenome of depression, and that one latent vector could be extracted from ruminating, depressive and anxiety symptoms, insomnia, and suicidal behaviors. In contrast, previous research examined rumination as a predictor (Riihimäki et al., 2016; Whisman et al., 2020), mediator (Lyon et al., 2021; Vidal-Arenas et al., 2022), and consequence (Whisman et al., 2020) of depression. In a longitudinal study, Whisman and colleagues (2020) discovered that rumination has a bidirectional association with changes in depressive symptoms in community adults, both as a risk factor and a consequence. Ruminating is a complete and partial mediator between depression and anxiety in adolescents and adults, respectively, and may increase the risk of comorbidity between these affective disorders (McLaughlin and Nolen-Hoeksema, 2011).

Our PLS model, however, shows that brooding (assessed as a trait) is a manifestation of the phenome of depression, which therefore should be regarded as the cause of its manifestations, including increased brooding (assessed as a trait), the severity of depression and anxiety (assessed as state features over the last one/two weeks), insomnia (assessed as a state feature over the last two weeks), and suicidal behaviors (assessed as a state feature over the last month). According to Treynor and colleagues (2003), the RRS was created as a self-report of ruminations about depressive symptoms that partially correlate with BDI items. In actuality, the RRS scale items reveal self-ratings of rumination regarding depressive symptoms based on BDI items such as exhaustion, motivation, concentration difficulties, personal failings, and regret (Beck et al., 1996; Nolen-Hoeksema and Morrow, 1991). Overall, our study indicates that increased brooding about depressive symptoms should be considered a manifestation of the phenome of depression, just like anxiety, loss of interest, suicidal ideation, insomnia, and cognitive deficits in attention, executive functions, and memory (Maes and Almulla, 2023). Moreover, once present, brooding can exacerbate the symptoms of depression by interfering with cognitive processes and reward processing, thereby increasing the flow of negative information, and decreasing the capacity to engage in positive responses (Koster et al., 2011; Owens and Gibb, 2017; Rutherford et al., 2023).

### Brooding and neuroticism as manifestations of depression

The third major finding of this study is that our initial PLS model (figure 4) demonstrating that brooding mediates the effects of neuroticism on the depressive phenome, could not be validated. Previously, Roelofs et al. (2008) and Lyon et al. (2021) found that brooding and refection mediate the effects of neuroticism on depressive symptoms. In our study, however, no discriminatory validity could be established between neuroticism and brooding or between neuroticism and the depressive phenome, thereby excluding the possibility that brooding is a mediator. Next, we were able to incorporate neuroticism, brooding, and all phenome features into one single latent vector, indicating that neuroticism and brooding are manifestations of the same core, namely the depression phenome.

With respect to neuroticism, our findings are consistent with those of Jirakran et al. (2023), who demonstrated that a latent vector can be extracted from neuroticism (a personality trait) and state characteristics of the depression phenome in major depressed patients with a longer duration of illness. We found, like Jirakran et al. (2023), that neuroticism is the personality trait that is most strongly associated with depression, whereas agreeableness and consciousness (both decreased in depression) are less significant depression-associated characteristics. Numerous studies have documented an association between neuroticism and depression (e.g., Ho et al., 2022; Merino et al., 2016; Whisman et al., 2020). In cross-sectional and longitudinal studies, neuroticism has been associated with rumination and depressive symptoms. Moreover, du Pont et al. (2019) reported that rumination and neuroticism have a shared genetic basis.

Numerous psychological theories could, of course, explain the significant relationships between depression, neuroticism, and ruminating. Those with neuroticism and a more ruminative response style, for example, may have abnormalities in reward processes, working memory, and executive functions, which may fuel ruminations about the negative effects of depression and contribute to negative self-evaluations and self-reflections. In addition, this process may result in a greater retention of negative cognition, leading to a decline in problem-solving and interpersonal functioning. Nevertheless, we demonstrated previously that the phenome of depression is strongly influenced by a variety of biological pathways that could lead to neuro-affective toxicity by interfering with affective-cognitive neuronal circuits (Al-Hakeim et al., 2023a), and thus all phenotypic characteristics of depression. Interestingly, the same biomarkers that underlie the depressive phenome are also strongly associated with neuroticism (Maes et al., 2023a). These findings provide additional evidence that neuroticism is a component of the phenome of depression, which is caused by biological pathways. Future research should investigate whether the same biomarkers are also linked to rumination and brooding. Since rumination may be a trait-like cognitive response to distress (Nolen-Hoeksema, 1991), it is conceivable that the phenome of the acute phase of depression exacerbates this trait, thereby contributing causatively to the phenome of MDD.

### Limitation

This study should be interpreted in light of its limitations. Firstly, it would be essential to investigate rumination during the partially remitted and remitted phases of depression, as well as the preclinical phase preceding the onset of depression. The incorporation of depression due to Long-COVID may be an intervening factor (Al-Hakeim et al., 2023b). Nevertheless, we included a few students who had previously suffered from mild acute COVID-19 infection and omitted participants who had suffered from moderate to severe COVID-19 infection. Since Long-COVID develops in patients with critical COVID-19 infection (Al-Hakeim et al., 2023b), we minimized bias due to previous COVID-19 infection.

## Conclusions

Overall, this study supports the hypothesis that rumination and neuroticism (both assessed as traits) are manifestations of the depression phenome. The results of this study add to our understanding that brooding could be considered as an important manifestation of the depressive phenome. Therefore, brooding should be incorporated in rating scales that measure the severity of depression and probably also as a diagnostic criterion to make the diagnosis of depression. Future studies should examine whether the biological pathways, which may lead to neuro-affective toxicity in depression, are associated with brooding.

## Data Availability

The last author (MM) will reply to reasonable requests for the dataset used in the current study after it has been fully utilized by all authors.

## Ethical approval and consent to participate

The research project (IRB no.351/63) was approved by the Institutional Review Board of Chulalongkorn University’s institutional ethics board, Bangkok, Thailand, which is in compliance with the International Guideline for Human Research protection as required by the Declaration of Helsinki, The Belmont Report, CIOMS Guideline and International Conference on Harmonization in Good Clinical Practice (ICH-GCP).

## Author’s contributions

AV and MM carried out the current study’s design. The data was gathered by AV. The statistical evaluation was done by AV and MM. Each author contributed to the writing and rewriting of the work, and they have all given their consent for submission of the completed version.

## Acknowledgments

Not applicable.

## Electronic supplementary file (ESF)

**ESF, Table 1.** Description of rumination items from The Ruminative Response Scale (RRS)

| Items | Description |
| --- | --- |
| RRS1 | Think: how lonely you are. |
| RRS2 | Think: "I won't be able to do my job if I don't snap out of this" |
| RRS3 | Think: how exhausted and weak you feel. |
| RRS4 | Think: how difficult it is to focus. |
| RRS5 | Think: "What have I done to deserve this?" |
| RRS6 | Think: how helpless and unmotivated you feel. |
| RRS7 | Analyze recent occurrences to try to figure out why you're depressed. |
| RRS8 | Think: how you seem not to be feeling anything anymore. |
| RRS9 | Think: "Why can't I get started?" |
| RRS10 | Think: "Why do I always react like this?" |
| RRS11 | Go away by yourself and reflect on why you feel this way. |
| RRS12 | Make a list of your thoughts and examine them. |
| RRS13 | Think: about recent circumstance in which you wish things had gone differently. |
| RRS14 | Think: "If I keep feeling like this, I won't be able to concentrate." |
| RRS15 | Think: "Why do I have problems that others do not?" |
| RRS16 | Think: "How come I can't handle things better?" |
| RRS17 | Think: how depressed you are. |
| RRS18 | Think: about all your weaknesses, foibles, faults, and errors. |
| RRS19 | Think: how you don't feel like undertaking anything. |
| RRS20 | Analyze your personality to try to figure out why you're depressed. |
| RRS21 | Go somewhere alone to reflect on your thoughts. |
| RRS22 | Think: how furious you are with yourself. |

**ESF, Table 2.** Descriptions of suicide items from The Columbia–Suicide Severity Rating Scale (C-SSRS)

**ESF, Table 2.**
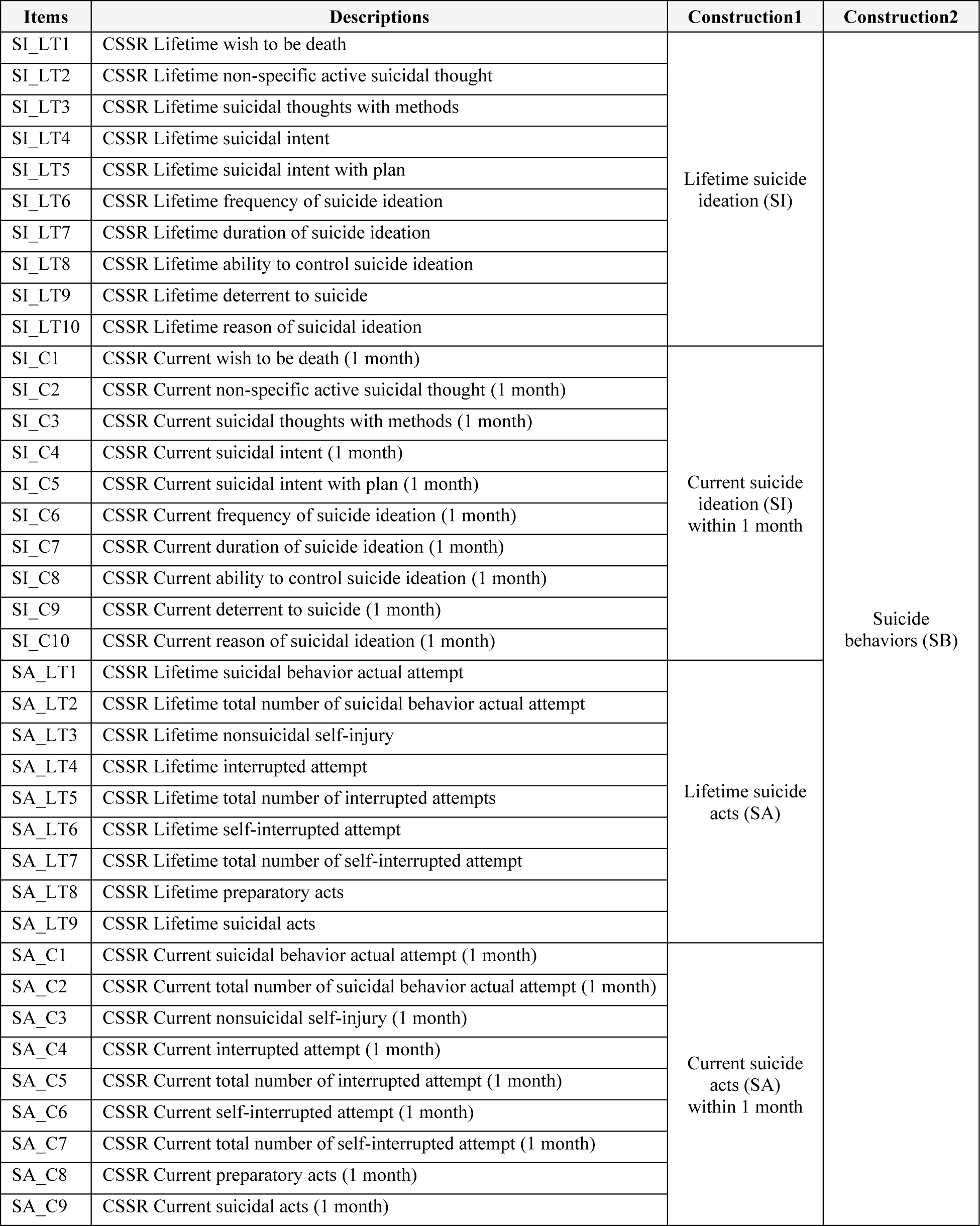
Descriptions of suicide items from The Columbia–Suicide Severity Rating Scale (C-SSRS)

| Items | Descriptions | Construction1 | Construction2 |
| --- | --- | --- | --- |
| SI_LT1 | CSSR Lifetime wish to be death | Lifetime suicide ideation (SI) | Suicide behaviors (SB) |
| SI_LT2 | CSSR Lifetime non-specific active suicidal thought |  |  |
| SI_LT3 | CSSR Lifetime suicidal thoughts with methods |  |  |
| SI_LT4 | CSSR Lifetime suicidal intent |  |  |
| SI_LT5 | CSSR Lifetime suicidal intent with plan |  |  |
| SI_LT6 | CSSR Lifetime frequency of suicide ideation |  |  |
| SI_LT7 | CSSR Lifetime duration of suicide ideation |  |  |
| SI_LT8 | CSSR Lifetime ability to control suicide ideation |  |  |
| SI_LT9 | CSSR Lifetime deterrent to suicide |  |  |
| SI_LT10 | CSSR Lifetime reason of suicidal ideation |  |  |
| SI_C1 | CSSR Current wish to be death (1 month) | Current suicide ideation (SI) within 1 month |  |
| SI_C2 | CSSR Current non-specific active suicidal thought (1 month) |  |  |
| SI_C3 | CSSR Current suicidal thoughts with methods (1 month) |  |  |
| SI_C4 | CSSR Current suicidal intent (1 month) |  |  |
| SI_C5 | CSSR Current suicidal intent with plan (1 month) |  |  |
| SI_C6 | CSSR Current frequency of suicide ideation (1 month) |  |  |
| SI_C7 | CSSR Current duration of suicide ideation (1 month) |  |  |
| SI_C8 | CSSR Current ability to control suicide ideation (1 month) |  |  |
| SI_C9 | CSSR Current deterrent to suicide (1 month) |  |  |
| SI_C10 | CSSR Current reason of suicidal ideation (1 month) |  |  |
| SA_LT1 | CSSR Lifetime suicidal behavior actual attempt | Lifetime suicide acts (SA) |  |
| SA_LT2 | CSSR Lifetime total number of suicidal behavior actual attempt |  |  |
| SA_LT3 | CSSR Lifetime nonsuicidal self-injury |  |  |
| SA_LT4 | CSSR Lifetime interrupted attempt |  |  |
| SA_LT5 | CSSR Lifetime total number of interrupted attempts |  |  |
| SA_LT6 | CSSR Lifetime self-interrupted attempt |  |  |
| SA_LT7 | CSSR Lifetime total number of self-interrupted attempt |  |  |
| SA_LT8 | CSSR Lifetime preparatory acts |  |  |
| SA_LT9 | CSSR Lifetime suicidal acts |  |  |
| SA_C1 | CSSR Current suicidal behavior actual attempt (1 month) | Current suicide acts (SA) within 1 month |  |
| SA_C2 | CSSR Current total number of suicidal behavior actual attempt (1 month) |  |  |
| SA_C3 | CSSR Current nonsuicidal self-injury (1 month) |  |  |
| SA_C4 | CSSR Current interrupted attempt (1 month) |  |  |
| SA_C5 | CSSR Current total number of interrupted attempt (1 month) |  |  |
| SA_C6 | CSSR Current self-interrupted attempt (1 month) |  |  |
| SA_C7 | CSSR Current total number of self-interrupted attempt (1 month) |  |  |
| SA_C8 | CSSR Current preparatory acts (1 month) |  |  |
| SA_C9 | CSSR Current suicidal acts (1 month) |  |  |

**ESF, Table 3.** Principle component (PC) analyses of suicide

**ESF, Table 3.**
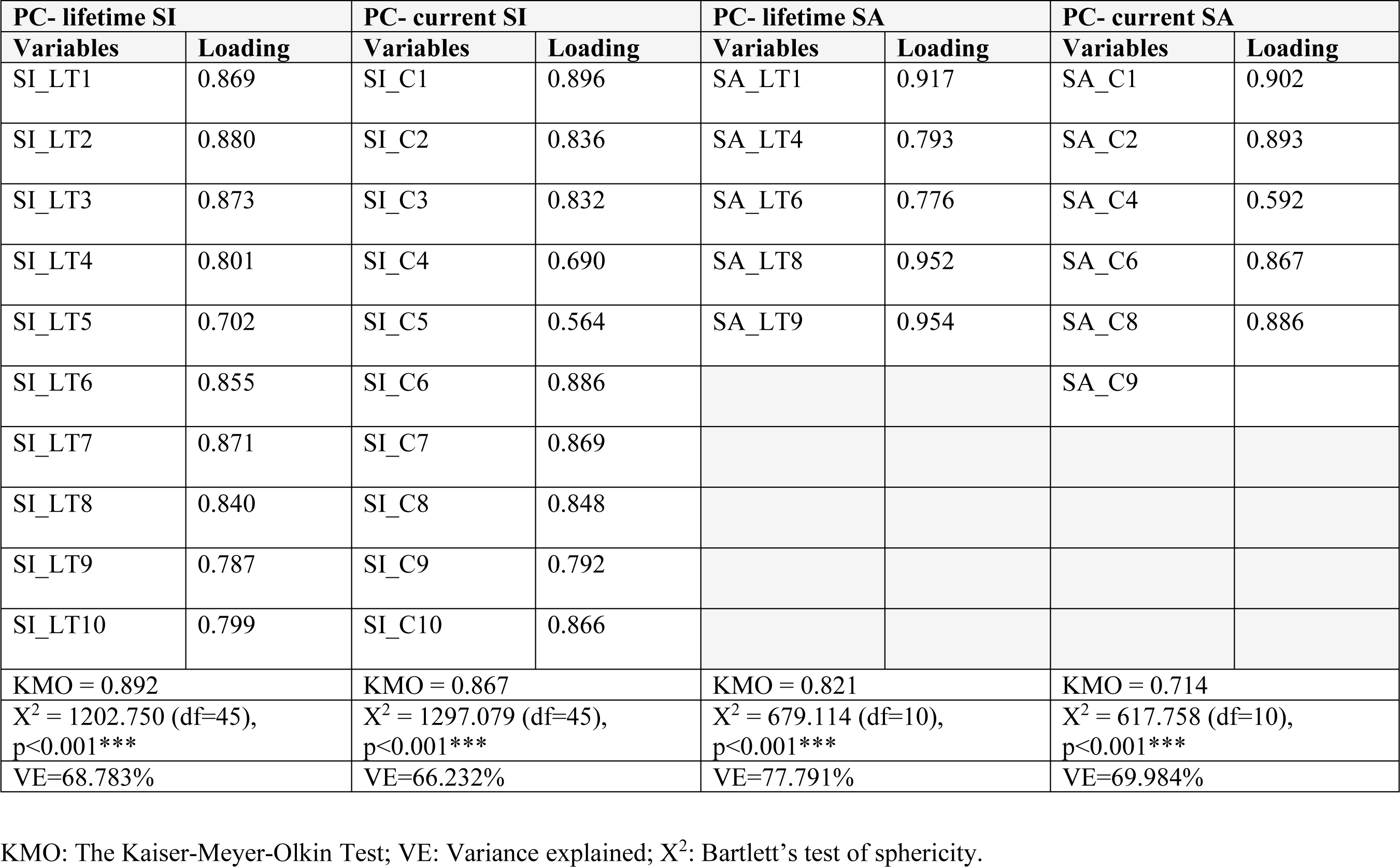
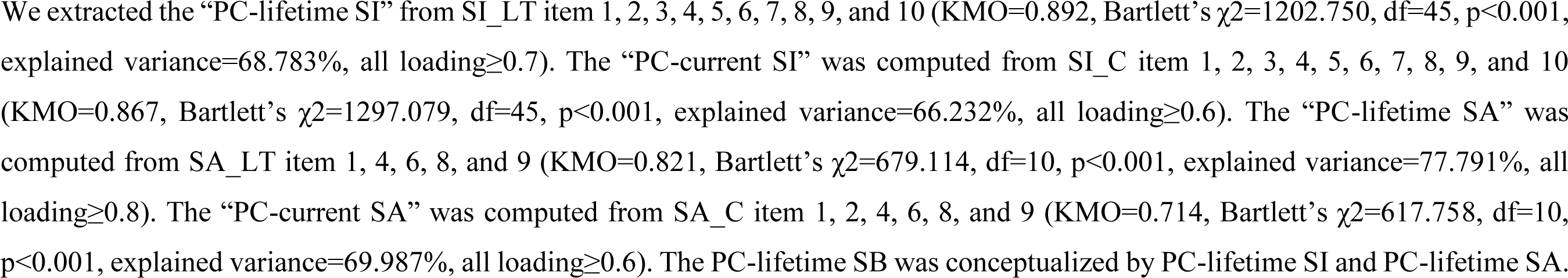
Principle component (PC) analyses of suicide

